# Immune phenotyping based on neutrophil-to-lymphocyte ratio and IgG predicts disease severity and outcome for patients with COVID-19

**DOI:** 10.1101/2020.03.12.20035048

**Authors:** Bicheng Zhang, Xiaoyang Zhou, Chengliang Zhu, Fan Feng, Yanru Qiu, Jia Feng, Qingzhu Jia, Qibin Song, Bo Zhu, Jun Wang

## Abstract

**Background:** A recently emerging respiratory disease named coronavirus disease 2019 (COVID-19) has quickly spread across the world. This disease is initiated by severe acute respiratory syndrome coronavirus-2 (SARS-CoV-2) and uncontrolled cytokine storm, but it remains unknown as to whether a robust antibody response is related to clinical deterioration and poor outcome in laboratory-confirmed COVID-19 patients.

**Methods:** Anti-SARS-CoV-2 IgG and IgM antibodies were determined by chemiluminescence analysis (CLIA) in COVID-19 patients from a single center in Wuhan. Median IgG and IgM levels in acute and convalescent-phase sera (within 35 days) for all included patients were calculated and compared among severe and nonsevere patients. Immune response phenotyping based on late IgG levels and neutrophil-to-lymphocyte ratio (NLR) was characterized to stratify patients with different disease severities and outcome. Laboratory parameters in patients with different immune response phenotypes and disease severities were analyzed.

**Findings:** A total of 222 patients were included in this study. IgG was first detected on day 4 of illness, and its peak levels occurred in the fourth week. Severe cases were more frequently found in patients with high IgG levels, compared to those who with low IgG levels (51.8% versus 32.3%; p=0.008). Severity rates for patients with NLR^hi^IgG^hi^, NLR^hi^IgG^lo^, NLR^lo^IgG^hi^, and NLR^lo^IgG^lo^ phenotype was 72.3%, 48.5%, 33.3%, and 15.6%, respectively (p<0.0001). Furthermore, severe patients with NLR^hi^IgG^hi^, NLR^hi^IgG^lo^ had higher proinflammatory cytokines levels including IL-2, IL-6 and IL-10, and decreased CD4+ T cell count compared to those with NLR^lo^IgG^lo^ phenotype (p<0.05). Recovery rate for severe patients with NLR^hi^IgG^hi^, NLR^hi^IgG^lo^, NLR^lo^IgG^hi^, and NLR^lo^IgG^lo^ phenotype was 58.8% (20/34), 68.8% (11/16), 80.0% (4/5), and 100% (12/12), respectively (p=0.0592). Dead cases only occurred in NLR^hi^IgG^hi^ and NLR^hi^IgG^lo^ phenotypes.

**Interpretation:** COVID-19 severity is associated with increased IgG response, and an immune response phenotyping based on late IgG response and NLR could act as a simple complementary tool to discriminate between severe and nonsevere COVID-19 patients, and further predict their clinical outcome.

**Research in context:** *Evidence before this study:* Following SARS-CoV-2 infection, a high viral load and overexuberant host immune response involving innate and acquired immunity, simultaneously contributes to the pathogenesis of COVID-19 and organ injury. Through searching PubMed and the China National knowledge infrastructure databases up to March 12, 2020, no published article focusing on anti-SARS-CoV-2 IgG-mediated immune response was identified.

*Added value of this study:* We evaluated antibody response within 35 days after symptom onset in laboratory-confirmed case with COVID-19 as one component of an overall exaggerated immune activation in severe SARS-CoV-2 infection, and developed an immune phenotyping based on late IgG response and NLR that could help determine disease severity and clinical outcome of COVID-19 patients. Severe cases were more frequently found in patients with high IgG levels, compared to those who with low IgG levels (51.8% versus 32.3%). Severity rates for patients with NLR^hi^IgG^hi^, NLR^hi^IgG^lo^, NLR^lo^IgG^hi^, and NLR^lo^IgG^lo^ phenotype was 72.3%, 48.5%, 33.3%, and 15.6%, respectively. Furthermore, severe patients with NLR^hi^IgG^hi^, NLR^hi^IgG^lo^ had higher proinflammatory cytokines levels including IL-2, IL-6 and IL-10, and decreased CD4+ T cell count compared to those with NLR^lo^IgG^lo^ phenotype. Recovery rate for severe patients with NLR^hi^IgG^hi^, NLR^hi^IgG^lo^, NLR^lo^IgG^hi^, and NLR^lo^IgG^lo^ phenotype was 58.8% (20/34), 68.8% (11/16), 80.0% (4/5), and 100% (12/12), respectively.

*Implications of all the available evidence:* COVID-19 severity is associated with a high viral load and overexuberant IgG response. We developed an immune response phenotyping based on NLR and IgG that could act as a simple complementary tool to discriminate between severe and nonsevere COVID-19 patients and would be helpful in guiding clinical decision.

## Introduction

Since December 2019, coronavirus disease 2019 (COVID-19) caused by severe acute respiratory syndrome coronavirus-2 (SARS-CoV-2) has quickly spread across the world.^1^ Approximately 20-30% of cases would develop severe illness, and some need further intervention in intensive care unit. Organ dysfunction including acute respiratory distress syndrome, shock, acute cardiac injury, and acute renal injury, could occur in severe cases with COVID-19, which lead to poor clinical outcome.^2,3^ Following SARS-CoV-2 infection, a high viral load and overexuberant host immune response involving innate and acquired immunity, simultaneously contributes to the pathogenesis of COVID-19 and organ injury.^2-4^ The activated host immunity is characterized as lymphopenia, cytokine release storm (CRS), and dysfunctional immune responses to virus-specific antigen. Increasing clinical data indicated that the neutrophil-to-lymphocyte ratio (NLR) was identified as a powerful predictive and prognostic indicator for severe COVID-19. However, the dynamic of anti-SARS-CoV-2 antibody upon virus infection and their relation to disease status and outcome remains to be determined.^5^

Here, we evaluated antibody response within 35 days after symptom onset in laboratory-confirmed case with COVID-19 as one component of an overall exaggerated immune activation in severe SARS-CoV-2 infection, and developed an immune phenotyping based on late IgG response and NLR that could help determine disease severity and clinical outcome of COVID-19 patients.

## Methods

### Data collection

All included patients with COVID-19 had been admitted to the Renmin Hospital of Wuhan University, from January 13, 2020 to March 1, 2020. A total of 222 laboratory-confirmed COVID-19 patients were included in this study. The confirmed diagnosis of COVID-19 was defined as a positive result by using real-time reverse-transcriptase polymerase-chain-reaction (RT-PCR) detection for routine nasal and pharyngeal swab specimens or anti-SARS-CoV-2 antibody assay. Serum samples were collected at admission or convalescent-phase and were dated from the day of initial symptom onset. We retrospectively evaluated their anti-SARS-CoV-2 antibody response, clinical disease severity, and clinical outcome. This study received approval from the Research Ethics Committee of the Renmin Hospital of Wuhan University, Wuhan, China (approval number: WDRY2020-K094). The Research Ethics Committee waived the requirement informed consent before the study started because of the urgent need to collect epidemiological and clinical data. We analyzed all the data anonymously.

The clinical features, including clinical symptoms, signs, laboratory analyses, treatment, and outcome, were obtained from the hospital’s electronic medical records according to previously designed standardized data collection forms. The date of symptom onset, initial diagnosis of COVID-19, and death were recorded accurately. To increase the accuracy of collected data, two researchers independently reviewed the data collection forms. We also directly communicated with patients or their family members to ascertain the epidemiological and symptom data.

### Antibody and cytokine assay

Anti-IgG and anti-IgM antibodies were detected using Human SARS-CoV-2 IgG and IgM Chemiluminescence Analysis (CLIA) Assays panel (Shenzhen YHLO Biotech Co.,Ltd., Shenzhen, China) and the high-speed CLIA system iFlash 3000 (Shenzhen YHLO Biotech Co.,Ltd., Shenzhen, China). Proinflammatory cytokines including interleukin (IL)-2, IL-4, IL-6, IL-10, interferon-γ (IFN-γ), and tumor necrosis factor-α (TNF-α) were detected using Human Cytokine Standard Assays panel (ET Healthcare, Inc., Shanghai, China) and the Bio-Plex 200 system (Bio-Rad, Hercules, CA, USA) according to the manufacturer’s instructions. NLR was calculated by dividing the absolute neutrophil count by the lymphocyte count.

### Statistical analysis

Descriptive analyses were used to determine the patients’ epidemiological and clinical features. Continuous variables were presented as median and interquartile range (IQR), and categorical variables were expressed as the percentages in different categories. Means for continuous variables were compared using independent group *t* tests when the data were normally distributed; otherwise, the Mann-Whitney test was used. The Chi-squared test or Fisher’s exact test was adopted for category variables. Statistical analyses in this study were performed with use of STATA 15.0 software (Stata Corporation, College Station, TX, USA). A two-sided *P* value less than 0.05 was considered statistically significant.

## Results

A total of 222 patients with a diagnosis of laboratory-confirmed COVID-19 recorded in the Renmin Hospital of Wuhan University were analyzed. Median age was 62 years (IQR; range from 52 to 69 years), and 48.2% of patients were male. 39.2% of patients were severe at the time of sampling. As of March 12, 2020, 5 patients (2.3%) died. A total of 121 patients (54.5%) required supplemental oxygen at some stage of illness. A total of 111 patients were administrated with high-dose corticosteroid. The number of patients receiving mechanical ventilation and administration of intravenous immunoglobin were 31 (14.0%) and 123 (55.4%), respectively. 194 patients recovered from this infected disease, and 59 severe patients recovered by anti-viral and supported therapy.

All patients had convalescent-phase sera for analysis. Of these, 98.6% of patients had anti-SARS-CoV-2-IgG detected in sera, and 82.0% had anti-SARS-CoV-2-IgM detected in sera. As shown in figure 1A, IgG was first detected on day 4 of illness, and its peak levels occurred in the fourth week, whereas IgM was first detected on day 3 of illness, and its peak levels occurred in the second week. Median IgG and IgM levels in convalescent-phase sera (within 35 days) for all included patients were compared among severe and nonsevere patients. Higher IgM levels were detected in patients with severe disease compared to those with nonsevere disease at early stage (<14 days), whereas higher IgG levels were detected at late stage (≥21 days) (figure 1B and 1C). We used median as cut-off value to stratify high and levels of IgM and IgG. Interestingly, severe cases were more frequently occurred in patients with low IgM levels (<34.1 AU/mL) than those with high IgM levels (≥3.04 AU/mL) (81.3% versus 40%; p=0.024) (figure 1D). Severe cases were more frequently found in patients with high IgG levels (≥116.9 AU/mL), compared to those who with low IgG levels (<116.9 AU/mL) (51.8% versus 32.3%; p=0.008) (figure 1E).

**Figure 1:**
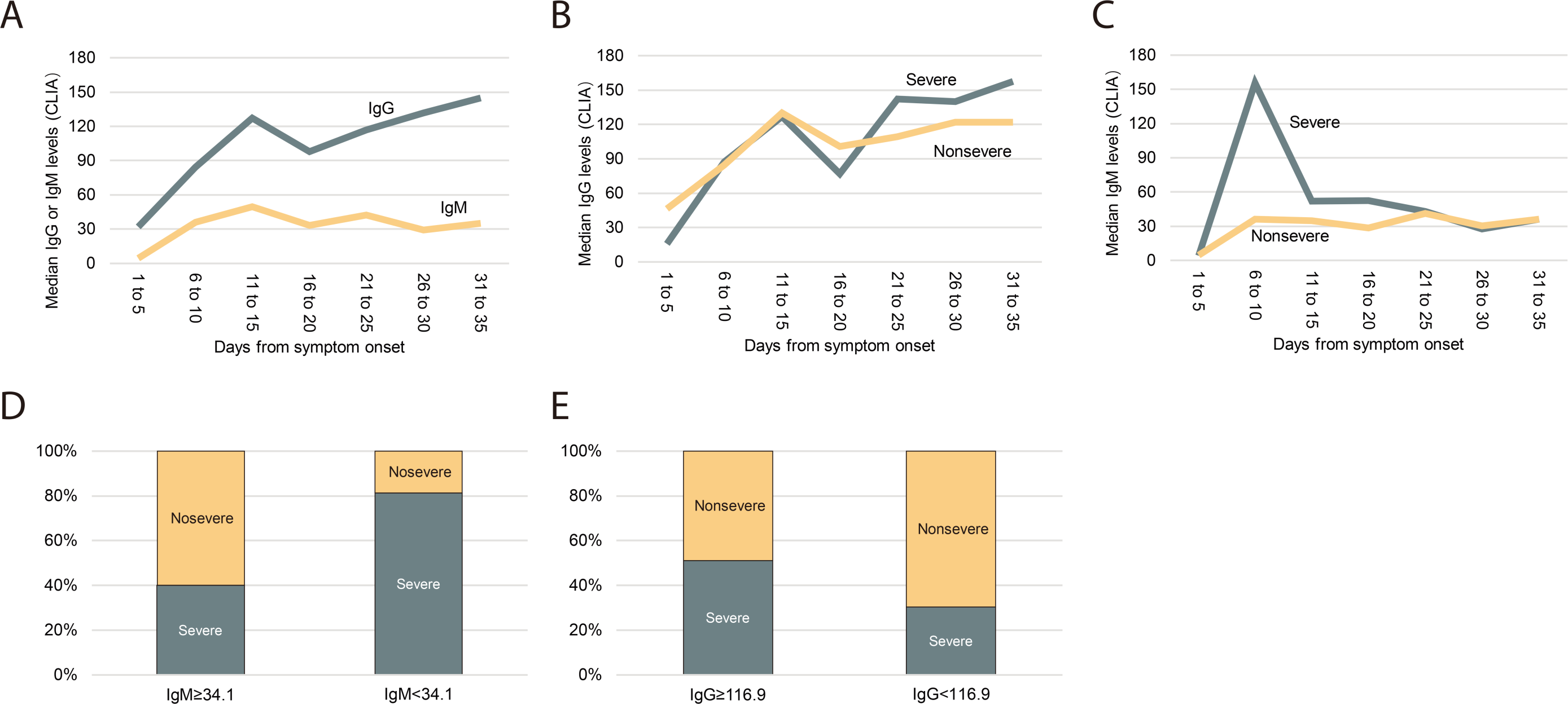
Median anti-SARS-CoV-2 IgG and IgM levels in patients with severe or nonsevere illness within 35 days after symptom onset. (A) Median IgG and IgM levels in all patients. (B) Comparing median IgG levels between severe and nonsevere patients. (C) Comparing median IgM levels between severe and nonsevere patients. (D) Comparing the frequency of severity and nonseverity between patients with low IgM levels (<34.1 AU/mL) or high IgM levels (≥3.04 AU/mL). (E) Comparing the frequency of severity and nonseverity between patients with low IgG levels (<116.9 AU/mL) or high IgG levels (≥116.9 AU/mL). CLIA: chemiluminescence analysis.

Considering NLR is linked to innate immunity, and anti-IgG response is an indicator of acquired immunity, we stratified patients at late stage into four different immune response phenotypes: high NLR and high IgG levels (NLR^hi^IgG^hi^), high NLR and low IgG levels (NLR^hi^IgG^lo^), low NLR and high IgG levels (NLR^lo^IgG^hi^), and low NLR and high IgG levels (NLR^lo^IgG^lo^), according to NLR (cutoff: 3.04) and detected IgG levels (cutoff: 116.9 AU/mL). Severity rates for patients with NLR^hi^IgG^hi^, NLR^hi^IgG^lo^, NLR^lo^IgG^hi^, and NLR^lo^IgG^lo^ phenotype were 72.3% (34/47), 48.5% (16/33), 33.3% (12/36), and 15.6% (5/32), respectively (p<0.0001) (figure 2A).

**Figure 2:**
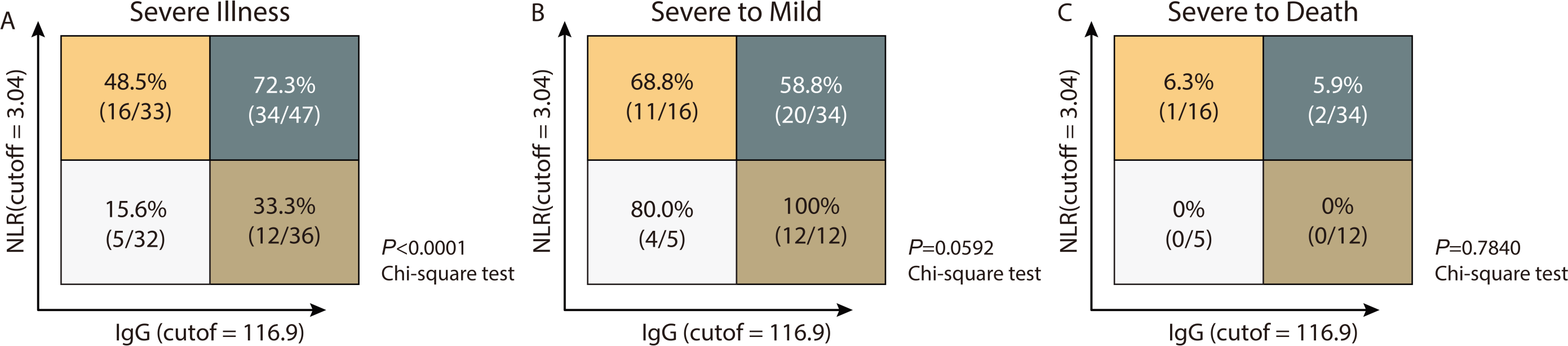
Immune response phenotyping with diverse disease severity according to NLR and IgG levels.

We next asked whether proinflammatory cytokines levels and T cell count were consistent with the above immune phenotyping. As shown in table 1, severe patients had higher proinflammatory cytokines levels including IL-2, IL-6 and IL-10 than nonsevere patients, especially in the NLR^hi^IgG^hi^ or NLR^hi^IgG^lo^ phenotype (p<0.05). Furthermore, severe patients with the NLR^hi^IgG^hi^ or NLR^hi^IgG^lo^ phenotype had higher proinflammatory cytokines levels including IL-2, IL-6 and IL-10, and decreased CD4+ T cell count, compared to those with the NLR^lo^IgG^lo^ phenotype (p<0.05). In particular, only IgG and IgM were higher in severe patients with the NLR^lo^IgG^hi^ phenotype than those in the NLR^lo^IgG^lo^ phenotype (p<0.05).

**Table 1:** Clinical characteristics, treatment and outcome of COVID-19 patients with different immune response phenotypes

| Patient groups | NLR<sup>high</sup>IgG<sup>low</sup> (n = 33) |  | NLR<sup>low</sup>IgG<sup>low</sup> (n = 32) |  | NLR<sup>high</sup>IgG<sup>high</sup> (n = 47) |  | NLR<sup>low</sup>IgG<sup>high</sup> (n = 36) |  |
| --- | --- | --- | --- | --- | --- | --- | --- | --- |
|  | Nonsevere (n = 17) | Severe (n = 16) | Nonsevere (n = 27) | Severe (n = 5) | Nonsevere (n = 13) | Severe (n = 34) | Nonsevere (n = 24) | Severe (n = 12) |
| IgG | 89.8 (55.3-103.9) | 87.3 (73.5-99.8) | 85.9 (74.6-93.7) | 87.1 (72.1-99.1) | 150.5 (138.0-172.3) | 160.3 (142.1-170.7)<sup>#</sup> | 158.7 (136.7-172.3) | 172.2 (151.9-188.2)<sup>#</sup> |
| IgM | 30.3 (9.8-81.2) | 68.3 (18.7-187.1) | 46.9 (23.2-107.3) | 31.5 (17.9-79.2) | 18.3 (8.9-30.6) | 34.1 (15.2-48.6) | 48.4 (24.9-94.0) | 35.3 (13.0-68.2)<sup>#</sup> |
| Age, years | 62 (47-78) | 62.5 (54.0-69.0) | 49 (37-58) | 48.0 (36-57) | 66.5 (56-73) | 70.5 (63.0-78.0)<sup>#</sup> | 59.5 (54.5-64) | 62.0 (52.0-74.5) |
| Male | 41.2 (7/17) | 75.0 (12/16) | 52.0 (14/27) | 60.0 (3/5) | 61.5 (8/13) | 50.0 (17/34) | 41.7 (10/24) | 50.0 (6/12) |
| Comorbidity | 58.8% (10/17) | 43.8 (7/16) | 25.9 (7/27) | 60.0 (3/5) | 76.9 (10/13) | 64.7 (22/34) | 41.7 (10/24) | 91.7 (11/12) |
| WBC, × 10<sup>9</sup>/L | 5.9 (4.1-7.4) | 10.0 (7.4-19.2)<sup>*#</sup> | 4.9 (4.4-6.2) | 6.6 (4.5-6.9) | 6.3 (5.6-8.9) | 8.8 (6.3-11.1) | 4.7 (4.3-6.8) | 5.6 (4.1-6.8) |
| NEU, × 10<sup>9</sup>/L | 3.8 (2.9-5.4) | 8.0 (5.3-17.2)<sup>*#</sup> | 2.8 (2.3-3.1) | 4.1 (2.4-4.3) | 4.8 (3.9-6.3) | 6.7 (5.1-10.0)<sup>*#</sup> | 2.8 (2.2-3.8) | 2.9 (2.3-4.2) |
| LYM, × 10<sup>9</sup>/L | 0.9 (0.5-1.0) | 1.1 (0.8-1.3)<sup>#</sup> | 1.6 (1.3-2.0) | 1.5 (1.3-1.7) | 1.2 (0.8-1.5) | 0.9 (0.7-1.1)<sup>#</sup> | 1.6 (1.2-2.2) | 1.9 (1.3-2.0) |
| NLR | 4.2 (3.4-8.7) | 7.9 (4.1-22.6)<sup>*</sup> | 1.7 (1.4-2.1) | 2.1 (1.9-2.5) | 3.9 (3.5-4.9) | 6.7 (5.0-10.4)<sup>*</sup> | 1.8 (1.4-2.2) | 2.1 (1.5-2.3) |
| CD3+ T cells, × 10<sup>12</sup>/L | 586 (428-716) | 453 (274-953) | 1141 (864-1315) | 869 (676-1803) | 629.5 (341.5-825.5) | 847.5 (399-733) | 976 (839-1505) | 1301 (886-1412) |
| CD4+ T cells, × 10<sup>12</sup>/L | 416 (305-474) | 273 (60-462) | 662 (493-736) | 614 (449-1318) | 387 (215-516) | 375.5 (269-450)<sup>#</sup> | 604 (484-805) | 755 (596-972) |
| CD8+ T cells, × 10<sup>12</sup>/L | 176.5 (127-189) | 129 (110-295) | 409 (283-556) | 281 (197-545) | 220.5 (116-294) | 187.5 (105-281) | 403 (268-486) | 311 (291-503) |
| IL-2, pg/ml | 3.3 (3.0-4.1) | 3.2 (3.1-3.5) | 3.3 (3.0-3.8) | 3.2 (3.1-3.3) | 3.4 (2.7-4.0) | 4.0 (3.3-4.3)<sup>*#</sup> | 3.9 (3.1-4.2) | 3.8 (3.8-4.0) |
| IL-4, pg/ml | 3.2 (2.9-4.3) | 3.3 (2.9-3.6) | 3.3 (3.0-4.2) | 3.2 (3.0-3.6) | 3.0 (2.8-3.6) | 3.1 (2.9-3.6) | 3.6 (3.2-4.2) | 3.6 (3.5-3.8) |
| IL-6, pg/ml | 5.7 (4.5-12.5) | 24.4 (10.2-97.6)<sup>*#</sup> | 6.2 (4.4-8.0) | 6.7 (5.5-7.2) | 7.4 (5.9-20.1) | 24.8 (16.6-53.0)<sup>*#</sup> | 6.9 (4.3-9.1) | 9.7 (6.2-15.3) |
| IL-10, pg/ml | 5.6 (4.2-7.6) | 8.0 (5.7-25.0) | 5.1 (4.7-5.8) | 5.4 (4.8-5.5) | 5.4 (4.4-6.6) | 9.0 (7.3-14.4)<sup>*#</sup> | 5.7 (4.4-7.3) | 4.8 (5.5-6.2) |
| TNF-α, pg/ml | 3.1 (2.9-3.3) | 3.2 (2.9-3.2) | 2.9 (2.7-3.3) | 3.3 (3.2-3.5) | 4.7 (3.3-6.4) | 3.8 (3.0-6.6) | 3.0 (2.8-3.9) | 5.0 (3.1-11.7) |
| IFN-γ, pg/ml | 3.6 (2.2-4.3) | 3.4 (2.7-4.4) | 3.4 (2.4-4.1) | 3.6 (2.9-4.0) | 4.2 (2.6-4.8) | 3.3 (3.0-4.6) | 3.8 (3.1-4.4) | 4 (3.9-4.6) |
| Oxygen therapy | 29.4 (5/17) | 100 (16/16) | 18.5 (5/27) | 100 (5/5) | 46.2 (6/13) | 94.1 (32/34) | 37.5 (9/24) | 75.0 (9/12) |
| Steroid | 35.3 (6/17) | 75.0 (12/16) | 40.7 (11//27) | 60.0 (3/5) | 76.9 (10/13) | 67.6 (23/34) | 37.5 (9/24) | 50.0 (6/12) |
| Mechanical<br>ventilation | 0 (0/17) | 43.8 (7/16) | 0 | 0 | 0 | 44.1 (15/34) | 0 (0/24) | 16.7 (2/12) |
| Recovery | 100 (19/19) | 68.8 (11/16) | 100 (29/29) | 80.0 (4/5) | 100 (16/16) | 58.8 (20/34) | 100 (28/28) | 100 (12/12) |

Data are presented as median (IQR), n/N (%), where N represents the total number of specific patients with available data. \*p<0.05 versus nonsevere within individual immune response phenotype; #p<0.05 versus severe in NLR<sup>low</sup>IgG<sup>low</sup> phenotype. NLR: neutrophil-to-lymphocyte ratio; WBC: white blood cell; NEU: neutrophil; LYM: lymphocyte; IL-2: interleukin-2; IL-4: interleukin-4; IL-6: interleukin-6; IL-10: interleukin-10; TNF-α : tumor necrosis factor-α ; IFN-γ : interferon-γ

We also analyzed the treatment and clinical outcome for patients with different combined immune response phenotypes. Follow up the patients so far, 44.1% of patients with NLR^hi^IgG^hi^ phenotype, 43.8% of patients NLR^hi^IgG^lo^ phenotype, 16.7% of patients with NLR^lo^IgG^hi^ phenotype received mechanical ventilation, but no patients with NLR^lo^IgG^lo^ phenotype were treated with mechanical ventilation. All nonsevere patients did not develop severe disease. Recovery rate for severe patients with NLR^hi^IgG^hi^, NLR^hi^IgG^lo^, NLR^lo^IgG^hi^, and NLR^lo^IgG^lo^ phenotype was 58.8% (20/34), 68.8% (11/16), 80.0% (4/5), and 100% (12/12), respectively (p=0.0592) (figure 2B). Dead cases only found in population with the NLR^hi^IgG^hi^ or the NLR^hi^IgG^lo^ phenotypes (figure 2C). However, too few severe cases at early stage had convalescent sera collected to allow separate analysis of anti-IgM response.

## Discussion

In this study, we found enhanced IgM levels at early stage, and high levels were more frequently found in patients with severe disease. IgG levels increased at late stage, whereas high levels of IgG were frequently found in patients with severe disease. These results indicated that beside the antiviral efficacy, the antibody response might be associated with secondary antibody-mediated organ damage. Using NLR and IgG levels detected in sera at late stage, we develop a combined immune response phenotype, which could predict disease severity and the outcome of COVID-19 patient. To our knowledge, this is the first in the literatures to combine indicators from innate and acquired immunity to predict disease severity and outcome.

Anti-SARS-CoV-2-IgG can be detected at day 4 after onset, and it has peak levels at the fourth week, which is similar to the Anti-SARS-CoV-IgG response profile.^6^ Previous data showed severe SARS was associated more robust serological responses including early seroconversion (<day 16) and higher IgG levels.^7^ Low antibody titres were observed in patients with mild SARS or diseases with other viral infection.^8-10^ Be consistent with findings, present study supported that a robust humoral response to SARS-CoV-2 correlated with disease severity. Furthermore, we believe that this robust acquired anti-IgG response not only contributes to viral clearance, but also leads to robust immune-mediated tissue damage.

Lymphopenia, neutrophilia, and high NLR are commonly presented and associated with more severe viral infection.^3,11,12^ Recent data indicated that the NLR was identified as a powerful predictive and prognostic factor for severe COVID-19.^5^ Thus, we develop simple combined immune response phenotypes using NLR, an indicator of innate immunity, and IgG, an indicator of acquired immunity. These four immune response phenotypes could be useful to distinguish the severe from the nonsevere cases and predict clinical outcome. We observed more severe cases occurred in NLR^hi^IgG^hi^ or NLR^hi^IgG^lo^ phenotypes, with only three dead cases. Patients with NLR^hi^IgG^hi^ or NLR^hi^IgG^lo^ appeared to be difficult to recover from severe SARS-CoV-2 infection, compared to NLR^lo^IgG^hi^ or NLR^lo^IgG^lo^ phenotype. A third of patients with NLR^lo^IgG^hi^ had critical illness, suggesting that IgG response alone could lead to disease deterioration. Furthermore, high Th1 cytokine IFN-γ, proinflammatory cytokines IL-6, TNF-α, and IL-10 were noted in NLR^hi^IgG^hi^ or NLR^hi^IgG^lo^ phenotypes. Accordingly, we speculated that anti-IgG humoral response directly contributed to tissue damage. In SARS-CoV/macaque models, Liu et al. found anti-spike IgG, the presence of high anti-spike IgG prior to viral clearance, abrogated wound-healing responses and promoted proinflammatory cytokines monocyte chemotactic protein 1 and IL-8 production and monocyte/macrophage recruitment and accumulation, eventually cause fatal acute lung injury during SARS-CoV infection.^13^

Thus, we proposed potential mechanisms associated with different immune response phenotypes and presented specific treatment recommendations which would be helpful in guiding clinical decision (figure 3). For example, in the NLR^hi^IgG^lo^ group, both virus directly mediated tissue damage and CRS are responsible for disease development. In this scenario, anti-virus, tocilizumab, and serum from convalescent should be considered. However, serum from convalescent should only be used in patients with low IgG levels and should not be used in the patients with high IgG level since its use might aggravate the disease especially in the patients with NLR^hi^IgG^hi^ phenotype.^14^ Tocilizumab treatment might not be beneficial for patients with low NLR, because these patients are in the immunosuppression stage rather than in CRS stage.^15^

**Figure 3:**
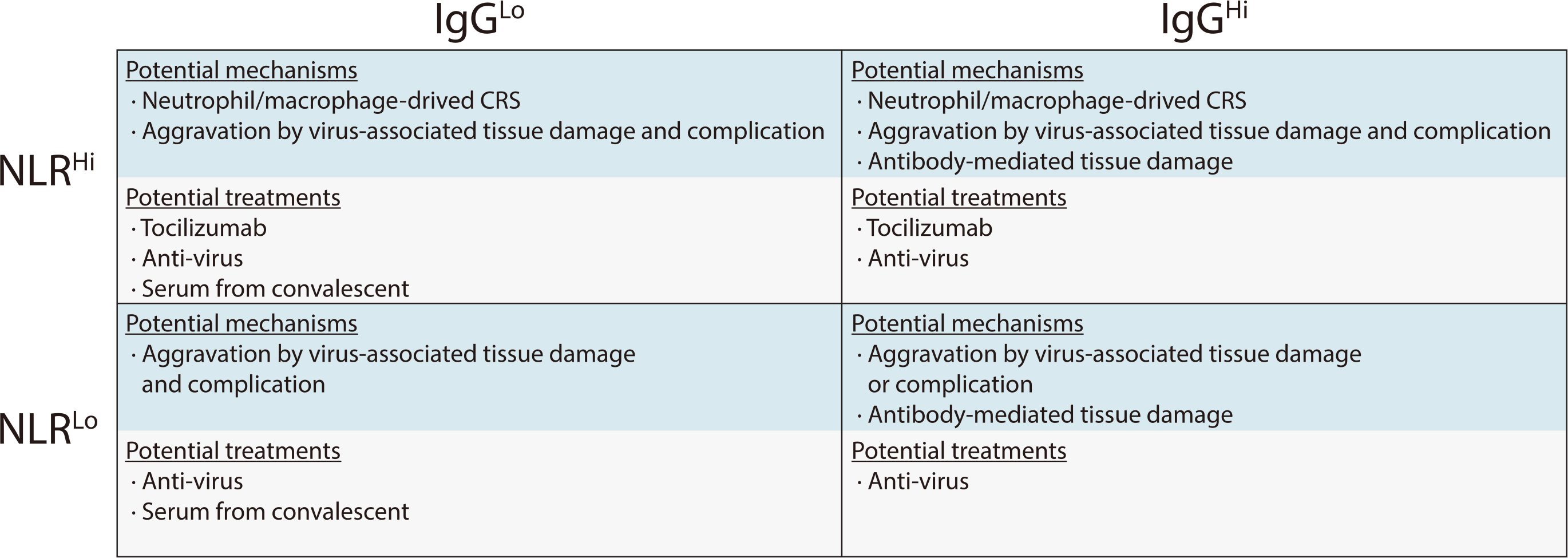
Immune response phenotyping with different immunological mechanisms associated with organ damage, and potential therapeutic strategies against severe COVID-19.

Our retrospective investigation had some limitations. Firstly, virus titers were not monitored during SARS-CoV-2 infection and patient recovery. Higher IgG levels, however, were previously detected in patients who had negative pre-discharge fecal RT-PCR results and SARS-CoV-infected rhesus macaques that had markedly reducing virus titers.^7^ Secondly, it is unknown whether the change or increase of IgM or IgA is related to disease severity. The mechanism responsible for the immunopathologic reaction of IgG remains elusive. Finally, the IgG response and its correlation to the severity of COVID-19 in patients without high-dose corticosteroid intervention have not been addressed. Nevertheless, our findings indicate that severe COVID-19 was associated with a more robust IgG response that can be developed as an acquired immunity-related marker to predictive disease severity, along with other innate immunity-relate makers such as NLR. Further study on the immunopathogenesis of SARS-CoV-2 infection is warranted.

## Data Availability

The data that support the findings of this study are available from the corresponding author on reasonable request. Participant data without names and identifiers will be made available after approval from the corresponding author and National Health Commission. After publication of study findings, the data will be available for others to request. The research team will provide an email address for communication once the data are approved to be shared with others. The proposal with detailed description of study objectives and statistical analysis plan will be needed for evaluation of the reasonability to request for our data. The corresponding author will make a decision based on these materials. Additional materials may also be required during the process.

## Contributors

JW, BZ and QS had the idea for and designed the study and had full access to all data in the study and take responsibility for the integrity of the data and the accuracy of the data analysis. BiZh, XZ and CZ contributed to writing of the report. BZ contributed to critical revision of the report. JW and BiZh contributed to the statistical analysis. All authors contributed to data acquisition, data analysis, or data interpretation, and reviewed and approved the final version.

## Acknowledgments

We acknowledge all health-care workers involved in the diagnosis and treatment of patients in Eastern Campus, Renmin Hospital of Wuhan University; we thank Prof Hong Zhou and Jiang Zheng for guidance in manuscript preparation.

